# Bioinformatics analyses and experimental validation of ferroptosis-related genes in bronchopulmonary dysplasia pathogenesis

**DOI:** 10.1101/2023.09.04.23295026

**Authors:** Yifan Luo, Zongli Zhang, Shibing Xi, Tao Li

## Abstract

**Objective:** We aimed to study the involvement of ferroptosis in the pathogenesis of bronchopulmonary dysplasia (BPD) by conducting bioinformatics analyses and identifying and validating the associated ferroptosis-related genes to explore new directions for treating BPD.

**Methods:** The dataset GSE32472 on BPD was downloaded from the public genome database. Using R language, differentially expressed genes (DEGs) between the BPD and normal group were screened. In the present study, we adopted weighted gene correlation network analysis (WGCNA) for identifying BPD-related gene modules and ferroptosis-related genes were extracted from FerrDb. Their results were intersected to obtain the hub genes. After that, to explore the hub gene-related signaling pathways, the hub genes were exposed to gene ontology enrichment analysis. With the purpose of verifying the mRNA expression of the hub genes, a single-gene gene set enrichment analysis and quantitative reverse transcription polymerase chain reaction were conducted. Immune cell infiltration in BPD was analyzed using the CIBERSORT inverse fold product algorithm.

**Results:** A total of 606 DEGs were screened. WGCNA provided the BPD-related gene module darkgreen4. The intersection of DEGs, intramodular genes, and ferroptosis-related genes revealed six ferroptosis-associated hub genes (*ACSL1*, *GALNT14*, *WIPI1*, *MAPK14*, *PROK2*, and *CREB5*). Receiver operating characteristic curve analysis demonstrated that the hub genes screened for BPD were of good diagnostic significance. According to the results of immune infiltration analysis, the proportions of a cluster of differentiation (CD)8, CD4 naive, and memory resting T cells and M2 macrophage were elevated in the normal group, and the proportions of M0 macrophage, resting mast cell, and neutrophils were increased in the BPD group.

**Conclusions:** A total of six ferroptosis-associated hub genes in BPD were identified in this study, and they may be potential new therapeutic targets for BPD.

## 1 Introduction

Bronchopulmonary dysplasia (BPD) refers to a commonly seen chronic respiratory disease in preterm infants, with its incidence showing negative relationship to gestational age and birth weight (1, 2). There is no specific treatment available for BPD. Most infants do not respond well to the present treatment. They thus are prone to develop permanent pulmonary sequelae such as emphysema and pulmonary hypertension and impaired physical and neurological development, imposing a huge economic burden on their families and society (3–7). The study of BPD pathogenesis can help provide timely intervention and treatment; thus, it is of great clinical value.

Ferroptosis represents a novel form of cell death that is distinct from apoptosis, necrosis, autophagy, as well as other forms of cell death. Usually, it can be regarded as a regulated form of cell death resulted from the combined effect of lipid peroxidation and iron ion accumulation. During ferroptosis, iron ions induce free radical production via catalytic reactions, leading to increased oxidative stress in the biological membranes and disruption of lipid metabolism, ultimately leading to cell death. Ferroptosis is tightly related to various disease pathogeneses, such as neurodegenerative diseases, tumors, and cardiovascular diseases related to the disorders of iron metabolism (8, 9). A study found reduced levels of glutathione peroxidase 4 and glutathione in the lung tissues of hyperoxia-treated neonatal rats and that using ferroptosis inhibitors reduced hyperoxia-induced lung injury (10). Therefore, targeted regulation of ferroptosis can be a new direction in treating BPD.

In the current work, we applied bioinformatics analyses to screen for ferroptosis-related differentially expressed genes (DEGs) between normal children and children with BPD. Additionally, we identified new targets for diagnosing and treating BPD and validated them via in vitro cellular experiments.

## 2 Materials and Methods

### 2.1 Microarray Data

Using “Bmnchopulmonary dysplasia” as the search term, the public genomics database (Gene Expression Omnibus [GEO]; http://www.ncbi.nlm.nih.gov/geo) provided the dataset. Then, the GSE32472 dataset was selected after further filtering using “Series” and “Homo sapiens” as constraints. The GPL6244 platform was used to generate the GSE32472 ([HuGene-1_0-st] Affymetrix Human Gene 1.0 ST Array [transcript (gene) version]). After removing the samples with no health status from the dataset, data from 294 blood samples from healthy children (n = 112) and children with BPD (n = 182) was obtained.

Ferroptosis-associated gene data were downloaded from the ferroptosis database (FerrDb; http://www.zhounan.org/ferrdb/current/; query date: April 3, 2023), and a total of 484 gene sets consisting of driver, suppressor, and marker genes were acquired after the assembly and deletion of duplicate values.

### 2.2 DEG Analysis

DEGs were screened with the use of the “limma” package in the R software, where the *p*-value of <0.05 and |fold change (FC)| > 1.3 could be applied to be the differential gene screening conditions. The screening results were then visualized in the form of volcano plots and heat maps.

### 2.3 Weighted Gene Co-Expression Network Analysis (WGCNA)

With gene expression profiles, this study calculated the median absolute deviation (MAD) of each gene, respectively. In addition, the top 50% of genes which had the smallest MAD value were eliminated. The correlation coefficients between genes were calculated using the WGCNA package, and the correlation coefficient matrix was transformed into a neighborhood matrix, whereby a gene co-expression network was constructed. With a topological overlap, the genes were clustered into different modules through the nearest neighbor measure, where different modules were grouped into their respective functional modules based on their intrinsic commonality and similarity. The settings were as follows: minimum module size, 30; sensitivity, 3; and a specific color was assigned to each module, where the grey module was regarded to be the gene collection that could not be assigned to any module. Using a dynamic shear tree approach, modules were identified. The module feature vectors were calculated to correlate with gene expression to obtain the module membership (MM), where key genes within modules were screened on the basis of |MM| > 0.8.

### 2.4 Hub Gene Screening and Identification

Venn diagrams were drawn with an online website (https://bioinformatics.psb.ugent.be/webtools/Venn/), and hub genes were obtained in line with the intersection of DEGs, modular genes acquired by WGCNA, and the ferroptosis-related gene sets. Through calculating the receiver operating characteristic (ROC) curve and measuring the area under the curve (AUC), the diagnostic ability of the screened hub genes could be assessed as biomarkers.

### 2.5 Enrichment Analysis

Functional enrichment analysis was adopted for investigating the underlying biological mechanisms of the hub gene effect on BPD. The gene ontology (GO) of these genes was analyzed. The final results were shown in the form of a chord diagram with the R software. Additionally, gene set enrichment analysis (GSEA) was conducted on the hub genes. The hub gene expression by the GSEA software was used to classify the samples into high-(≥50%) and low-(<50%) expression groups. Using a p-value of <0.05 and a false discovery rate of <0.1 as screening criteria, the data was downloaded from the molecular signatures database as c2.cp.kegg.v7.4.symbols.gmt subset for assessing the relevant pathways and molecular mechanisms.

### 2.6 Immune Infiltration and Immune-Related Factors

The relative proportions of 22 immune cell species in each sample in the GSE32472 dataset were quantified based on the CIBERSORT method. Pearson’s correlation analysis was also carried out for hub genes and immune cell infiltration.

### 2.7 Real Time Quantitative PCR (RT-qPCR)

The human lung adenocarcinoma epithelial cell line A549 was cultivated in Dulbecco’s modified Eagle medium including 10% fetal bovine serum (11995500, Gibco). The cells were divided into two groups, where one was treated at a high oxygen concentration (85%) and the other at an oxygen concentration of 21%. At 0, 12, 24, and 48 h following the treatment, the total RNA was extracted with TRIzol (DP424, TIANGEN), reverse transcription was conducted with the Reverse Transcription Kit (R122–01, Vazyme). The real-time quantitative polymerase chain reaction (RT-qPCR) was made with the Taq pro universal SYBR qPCR master mix (Q712–02, Vazyme). With β-actin being an internal reference, we adopted the 2^−ΔΔCt^ method for calculating the relative expression of target genes. The primers were designed at the National Center for Biotechnology Information website (https://www.ncbi.nlm.nih.gov/) and synthesized at Shanghai Bioengineering Co. Table 1 presents the primer sequences.

### 2.8 Cell Survival Rate

The cells were divided into control and model groups, inoculated in 96-well plates, 6 replicate wells were set up for each group of cells, 100ul of cell suspension was supplemented to each well, 21% oxygen stimulation was given to the control group, 85% oxygen stimulation was given to the model group, 10ul of CCK8 working solution was supplemented to each well after 48h, incubated in the incubator for 1h, and the absorbance value at 450nm was measured with the use of enzyme marker.

### 2.9 Statistical Analysis

Based on R software, we carried out all statistical analyses. Student’s *t*-tests were adopted for analyzing the significance of differential expression of ferroptosis-related genes in the GEO dataset. In order to compare gene expression between samples in cellular experiments, one-way analysis of variance was adopted. Pearson’s correlation analysis was employed to analyze the relationship between hub genes and immune cell infiltration. In addition, the *p*-value of <0.05 was thought to be of statistical significance.

## 3 Results

### 3.1 Onset of Ferroptosis in a Hyperoxia-Induced BPD Model

In a previous experiment in which we constructed an in vitro model of BPD using hyperoxia in A549 epithelial cells, we found that a large number of cells died after 48 h of hyperoxia stimulation, and also observed uneven distribution of cells with different morphology and size (Figs 1A and 1B). Combined with the previous literature reporting that the use of iron death inhibitors could reduce hyperoxia-induced lung tissue injury, it was speculated that hyperoxia could induce ferroptosis to promote the occurrence and development of BPD. And by RT-qPCR assay, we decreased mRNA expression of two each classical ferroptosis marker glutathione peroxidase 4 (GPX4) and increased expression levels of prostaglandin-endoperoxide synthase 2 (PTGS2) were detected in cells after 48 h of hyperoxia treatment with *p*-value < 0.01. (Fig 1C).

### 3.2 Ferroptosis-Related DEGs in GSE32472

To further investigate the role of ferroptosis in BPD, bioinformatics techniques were used to screen for ferroptosis-related genes in BPD.Totally 606 DEGs were acquired from the GSE32472 dataset, containing 354 upregulated and 252 downregulated genes (Figs 2A and 2B). With the soft threshold power being at 10, the independence scale reached 0.86, and the average linkage value was 22.52 (Figs 3B and 3C). Totally 27 different co-expression modules were acquired through dynamic tree cutting with the cut height being set to 0.25, and the minimum module size being set to 30 (Fig 3E). The modules were then analyzed for correlation with clinical features. The darkgreen4 module was highly related to BPD (r = 0.40, *p* = 7.9e−13, Fig 3D), indicating that genes in this module may be associated with a relevant biological function. Additionally, genomic and BPD phenotypes were significantly correlated in the darkgreen4 module (r = 0.75, *p* = 2.0e−107, Fig 3F). With the module membership (MM) > 0.8 and gene significance (GS) > 0.2 being screening conditions, totally 187 genes were obtained from the darkgreen4 module. These 187 genes were crossed with 606 DEGs and 484 ferroptosis-related genes, and finally, six hub genes were obtained (Fig 4A). Violin plots showed that in the GSE32472 dataset, the expression of all six genes was discovered to be reduced in the BPD group in relative to the control group (Fig 5).

### 3.3 Cellular experimental Validation of Hub Genes

To demonstrate the mRNA expression of four hub genes, RT-qPCR was carried out. The findings demonstrated reduced expression of *ACSL1*, *GALNT14*, *WIPI1,* and *MAPK14* in the hyperoxia model group in relative to the blank control group, and *p*-value< 0.05 (Fig 6).

### 3.4 Functional enrichment Analysis of Hub Genes

To explain the hub gene-associated biological functions and pathways, GO enrichment analysis was made (Fig 4B). According to the results, three of the six ferroptosis-related differential hub genes were involved in “mitogen-activated protein kinase (MAPK) p38 binding,” “nuclear factor of activated T cell protein binding,” “medium-chain fatty acid-coenzyme ligase activity,” “decanoic acid coenzyme ligase activity,” and “acyl coenzyme ligase activity,” “MAPK activity,” “long-chain fatty acid-coenzyme ligase activity,” “fatty acid ligase activity,” “polypeptide n-acetyl galactosyltransferase activity,” and other functions.

Single-gene GSEA–Kyoto encyclopedia of genes and genomes pathway analysis was adopted for assessing the correlation of BPD with hub genes. Figs 7A–7E illustrate the five major pathways enriched for each hub gene. Of these, ACSL1 was related to the peroxisome proliferator-activated receptor (PPAR) signaling pathway, antigen processing and delivery, shear bodies, complement and coagulation cascades, and the insulin signaling pathway. CREB5 was related to the gonadotropin-releasing hormone signaling pathway, dorsal–ventral axis formation, MAPK signaling pathway, regulation of the actin cytoskeleton, and ribosomes. GALNT14 was associated with ribosomes, autoimmune thyroid disease, cell adhesion molecules, cysteine MAPK14 (related to type 2 diabetes), glycerophospholipid metabolism, PPAR signaling pathway, transendothelial migration of leukocytes, and actin cytoskeleton regulation. PROK2 was related to pyruvate metabolism, autoimmune thyroid disease, N-glycan biosynthesis, valine, leucine, and isoleucine degradation, and antigen processing and delivery. WIPI1 was associated with glycerophospholipid metabolism, sucrose, and starch metabolism, epithelial cell signaling in *Helicobacter pylori* infection, endocytosis, and cancer pathways.

### 3.5 ROC Curve Analysis for Hub Genes

To build regression models on the basis of six marker genes, the glmnet package in the R software was applied. ROC curves showed that the model was able to distinguish between the control and BPD group samples (AUC = 0.787, Fig 8A). In addition, ROC curves were adopted for assessing the correlation between individual genes and BPD, with each gene having an AUC value of >0.6 (Fig 8B). The findings demonstrated that in relative to the individual marker genes, the logistic regression model showed a higher level of specificity and accuracy.

### 3.6 Immuno-Infiltration Analysis

The microenvironment is consisted of immune cells, extracellular matrix, and inflammatory and diverse growth factors and has vital implications for clinical treatment sensitivity and disease diagnosis. Using the CIBERSORT algorithm, the naïve proportions of 22 immune cells across the 112 control samples and 182 BPD samples were estimated and are presented in the bar and violin plots (Figs 9A, 9B). Compared with the control samples, the BPD samples showed increased proportions of M0 macrophages, resting type mast cells, and neutrophils. In contrast, a cluster of differentiation (CD)8, Cnaïveive and memory resting T cells, and M2 macrophage proportions were reduced. According to Pearson’s correlation analysis, there existed significant negative and positive correlations between ACSL1, GALNT14, WIPI1, MAPK14, PROK2, CREB5 and CD8 T cells, CD4-naive T cells, CD4 memory T cells, M0 macrophage, and neutrophils (Fig 10).

## 4 Discussion

Novel BPD is a disease that primarily presents with impaired alveolar capillary development, alveolar enlargement and simplification, increased interstitial fibrosis, pulmonary vascular abnormalities, and reduced branching (11). The pathogenesis of this disease is not completely understood. However, some studies have indicated that oxidative stress refers to one of the important mechanisms in the pathogenesis of BPD (12–14). Long-term hyperoxia exposure induces oxidative stress, which affects alveolar development and angiogenesis. Furthermore, oxidative stress leads to the accumulation of oxygen radicals and other reactive oxygen products in lung cells, thereby damaging cell structure and leading to cell death and, ultimately, BPD (15, 16). Therefore, inhibiting the oxidative/antioxidative imbalance in BPD and decreasing cell death may be a new therapeutic direction in the future.

As a recently found form of cell death, ferroptosis is featured by an iron-dependent increase in lipid peroxidation because of reactive oxygen species (17). Iron is widely present in life and is one of the elements essential for cell metabolism and growth. However, when there is an excess of iron ions in cells, large amounts of oxygen radicals and cytotoxic reactions are generated, which leads to cell damage and death. There existed a decrease in the number of cells in the model group with variable changes in cell morphology during the construction of a hyperoxic BPD cell model, and also detected a decrease in GPX4 mRNA expression and an upregulation of PTGS2 expression in the model group, which has not been reported before. In addition, Cross et al. (18, 19) reported that free iron concentrations in cord blood were significantly higher in preterm infants than in full-term infants and adults. Patel et al. (20) reported that increased cumulative iron supplementation in very low birth weight infants is an independent risk factor for BPD. Moreover, BPD is most commonly observed in preterm infants and is a crucial factor in preterm mortality and poor long-term prognosis. We therefore hypothesize that abnormal accumulation of iron in preterm infants may induce the onset of ferroptosis thereby promoting the development of BPD.

On this basis, this study further clarifies the function of ferroptosis in the development of BPD. 606 DEGs of BPD were obtained from the GEO database, disease key modules were acquired using WGCNA analysis, 197 differential genes within modules were obtained after the screening, and 484 ferroptosis-related genes were acquired from the FerrDb database. There were intersected to finally obtain 6 hub genes related to ferroptosis in BPD, which included the following: *ACSL1*, *GALNT14*, *WIPI1*, *MAPK14*, *PROK2*, and *CREB5*.

The ACSL family is a vital enzyme in fatty acid synthesis and catabolism. ACSL1 is mainly distributed in adipose and muscle tissues in humans and makes a crucial role in cellular energy metabolism and fatty acid metabolism. Zhang et al. (21) reported that ACSL1 decreased intracellular lipid peroxidation levels and resisted the development of ferroptosis during ovarian cancer metastasis. In this study, bioinformatics analyses revealed that ACSL1 as a ferroptosis gene was decreased in samples from the BPD group of the GEO dataset. qPCR validated the results to be consistent; therefore, we hypothesized that in children with BPD, downregulated ACSL1 promotes ferroptosis and participates in disease progression. GALNT14 is a member of the peptide N-acetylgalactosaminyltransferase (ppGalNAc-Ts). These enzymes can catalyze the transfer of N-acetyl-D-galactosamine (GalNAc) to hydroxyl groups on Serine and Threonine in target peptides, initiating protein O-glycosylation and affecting tumorigenesis and progression (22). Li et al. (23) reported that the downregulation of GALNT14 induced cellular ferroptosis by inhibiting mTOR activity. In this study, a BPD model was constructed using hyperoxia-stimulated A549 cells, and qPCR was performed to determine whether GALNT14 expression gradually decreased with the prolongation of hyperoxia stimulation, conforming to the trend of microarray analysis, indicating that GALNT14 may be engaged in BPD development through the induction of ferroptosis.WIPI1 is a protein involved in the process of cellular autophagy and is a member of the cofactor protein family. The main role of WIPI1 is to interact with PI3K-III during the formation of autophagosomal membranes and to assist in their localization to the initiation site of autophagosomal membranes, which triggers the autophagic process (24). Deneubourg et al. (25) proposed that in Parkinson’s disease, Huntington’s disease, and other neurodegenerative diseases, the deletion of autophagy-related genes leads to the accumulation of abnormal protein and autophagic vesicles, disrupting neuronal homeostasis and ultimately leading to cell death. In this study, we screened WIPI1 as a hub gene in BPD by microarray analysis and found that WIPI1 expression was notably reduced in lung epithelial cells after hyperoxia treatment; as a result, we speculate that WIPI1 may be involved in cell death in BPD. MAPK14 is a serine–threonine protein kinase which is a member of the p38MAPK subfamily. MAPK14 makes a vital regulatory role in various diseases, including pneumonia and acute lung injury, and inhibition of this pathway can alleviate inflammatory responses and oxidative stress levels in disease states (26). Furthermore, the p38 MAPK signaling pathway is also engaged in regulating various angiogenic regulators, including VEGF, bFGF, and PDGF, affecting the proliferation, migration, and angiogenic capacity of endothelial cells (27, 28). This study focused on constructing a BPD cell model and found that MAPK14 expression reduced at 12 and 24 h of hyperoxia stimulation but returned to unstimulated levels after 48 h of stimulation, suggesting that MAPK14 may inhibit the oxidative stress response in vivo at the early stage of BPD, but gradually increased as the organism was in an uncompensated state.PROK2, a member of the prokineticin family, is a neuropeptide hormone that is involved as a cytokine in diverse physiological and pathological processes, including reproductive and neurodevelopment, angiogenesis, immune response, and inflammatory response. In the circulatory system, PROK2 is involved in regulating angiogenesis and vascular function, and regulates blood pressure and cardiac function (29). Bao et al. (30) reported that treating primary neuronal cells with the ferroptosis inducer Erastin increased PROK2 expression and protected mitochondrial function. Furthermore, PROK2 is mainly involved in activating and chemotaxis of monocytes and T cells in immune and inflammatory responses (31, 32). Wang et al. (33) identified a neutrophil subtype with high PROK2 expression in the lung vessels of ALI mice, which exhibited an immune-activated state and could resist pathogens. CREB5 belongs to a family of CRE (cAMP response element) binding proteins that encode proteins that bind CRE specifically to c-Jun or CRE-BP1 as homodimers or heterodimers and function as CRE-dependent trans-activators, regulating metabolism, cell cycle, growth factors, immune regulation, reproductive development, signal transduction, and other related biological processes (34–36). Gu et al. (37) reported that AG1296 increased CREB5 expression in PAH lung vessels and promoted angiogenesis. Based on the above studies, it is hypothesized that ACSL1, GALNT14, WIPI1, MAPK14, PROK2, and CREB5 may be involved in BPD progression by mediating cell death, oxidative stress, or regulating angiogenesis. However, the specific mechanisms between these genes and BPD require further investigation.

Furthermore, as the immune system is not completely developed in preterm infants, exposure to various risk factors, such as prenatal and postnatal infections, hyperoxia, and mechanical ventilation, leads to oxidative stress in the functionally immature lungs, increased expression of proinflammatory cytokines, recruitment of neutrophils as the first line of immune defense, and subsequent release of chemokines to recruit monocytes to adhere to endothelial cells and promote their extravasation and differentiation into macrophages and ultimately compromise epithelial and vascular function (38). Furthermore, mast cells can disrupt angiogenesis in rat and mouse models of hyperoxia stress (39–42). However, prolonged exposure to hyperoxia reduces the number of CD4+ and CD8+ T cells, which results in defective T cell development in the thymus (43). In this study, the number of T cells CD8 and CD4 and macrophages M2 in BPD was found to be less than the control group. By the CIBERSORT method, the number of macrophages M0, mast cells, and neutrophils was higher than the control group. Furthermore, we found that ACSL1, GALNT14, WIPI1, MAPK14, PROK2, and CREB5 were associated with these immune cells.

This study has some limitations that should be addressed. We selected only one dataset for analysis and validated the characteristics of the hub ferroptosis-related genes in an external dataset. Owing to the difficulties in obtaining samples, we only performed *in vitro* cellular experiments to validate these genes, which will be further validated using animal models. Therefore, further corroboration in clinical practice is still required.

## 5 Conclusions

Through a bioinformatics approach, we identified six hub genes associated with ferroptosis in BPD (*ACSL1*, *GALNT14*, *WIPI1*, *MAPK14*, *PROK2*, and *CREB5*), which were additionally found to be associated with immune cells. Moreover, our findings offer a novel idea for studying the pathogenesis of BPD.

## Data Availability

All data are available from the GEO database (https://www.ncbi.nlm.nih.gov/geo/query/acc.cgi?acc=GSE32472)

## Acknowledgments

We thank all authors for their contributions to this research

## Supporting information

**S1 Fig. Hyperoxia promotes BPD by inducing ferroptosis.** (A) Morphological images of A549 cells. (B) CCK8 method to detect the effect of hyperoxia stimulation for 48h on the viability of A549 cells. (C) Detection of GPX4, PTGS2 mRNA expression by RT-qPCR after hyperoxia (oxygen concentration of 85%) stimulation of A549 cells for 48h.**P < 0.01, ***P < 0.001, compared to the control (oxygen concentration of 21%) group.

**S2 Fig. Identification of DEGs.** (A) The volcano plot of DEGs between the BPD and control groups in the GSE32472 dataset. (B) The heatmap of the top 20 DEGs between the BPD and control groups in the GSE32472 dataset.

**S3 Fig. Identification of key module genes in WGCNA.** (A) Sample clustering dendrogram. (B) The analysis of scale-free fit index for different soft threshold cases. (C) The analysis of average connectivity for different soft thresholds. (D) Heatmap of the relationship between gene modules and sample information; the various colors represent the different modules. (E) Gene clustering tree diagram based on the co-expression networks constructed with optimal soft thresholds after classifying genes into different modules. (F) Scatter plot of GS and MM correlations for darkgreen4 module.

**S4 Fig. Identification of the ferroptosis-related DEGs genes and GO enrichment analyses of hub genes.** (A) Venn diagram of all DEGs, darkgreen4 module genes in WGCNA, and ferroptosis genes from the FerrDb database. (B) Circle diagram for GO enrichment analysis of hub genes.

**S5 Fig. Violin plot showing the levels of six hub genes in BPD and control samples of the dataset GSE32472.** (A) ACSL1. (B)GALNT14. (C) WIPI1. (D) MAPK14. (E) PROK2. (F)CREB5

**S6 Fig. Validation of mRNA expression of selected hub genes.** (A)ACSL1.(B)GALNT14. (C) WIPI1. (D) MAPK14.where\**P* < 0.05, \*\**P* < 0.01, \*\*\**P* < 0.001, when compared with the control (0 h) group.

**S7 Fig. Single-gene GSEA enrichment analysis**.(A) ACSL1.(B)GALNT14. (C) WIPI1. (D) MAPK14. (E) PROK2. (F) CREB5.Note: six major pathways were selected for each gene.

**S8 Fig. ROC curve analysis of 6 hub genes.** (A) Evaluation of the logistic regression models for joint diagnostic markers to identify disease samples in the GSE32472 dataset. (B) Evaluation of 6 hub genes to identify the disease samples in the GSE32472 dataset.

**S9 Fig. Immune infiltration analysis.** (A) Bar chart showing the proportion of 22 immune cell types in each sample. (B) Expression of different immune cells between the BPD and control groups, where\**P* < 0.05, \*\**P* < 0.01, \*\*\**P* < 0.001, \*\*\*\**P* < 0.0001, compared with the control group.

**S10 Fig. Correlation between the six hub genes and immune infiltration.**

**S1 Table.**
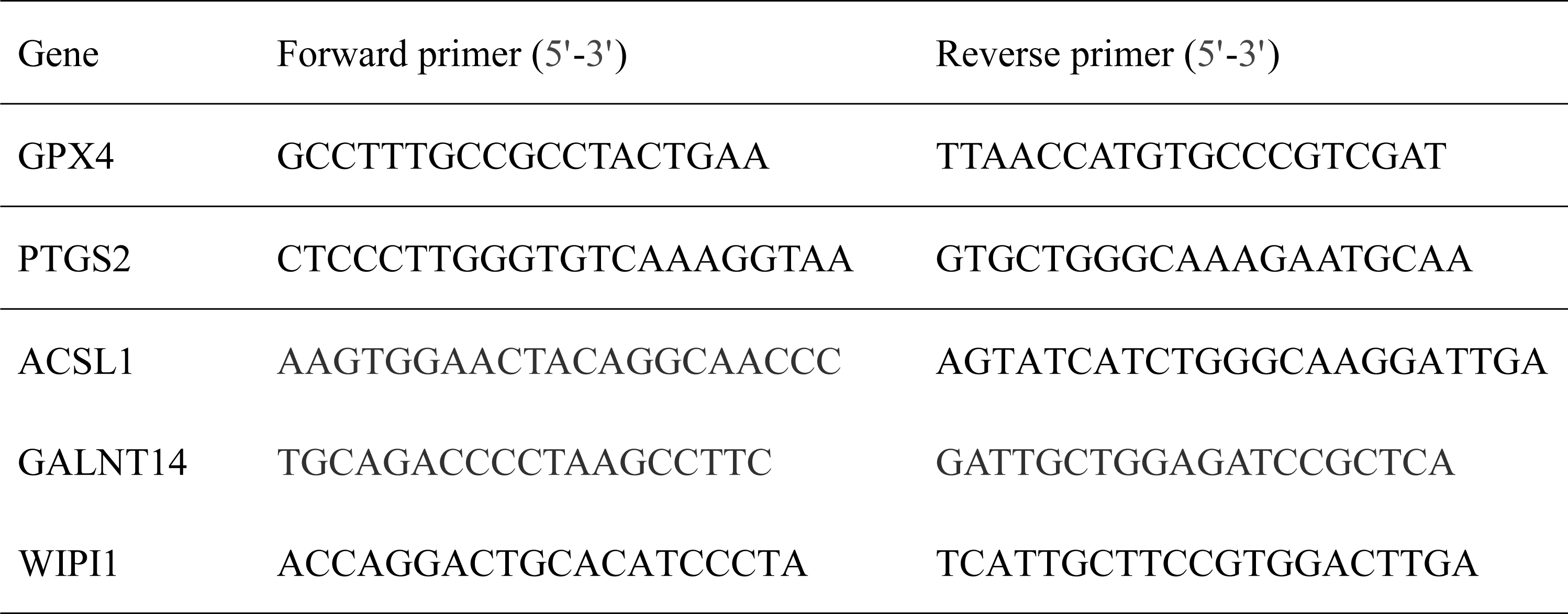
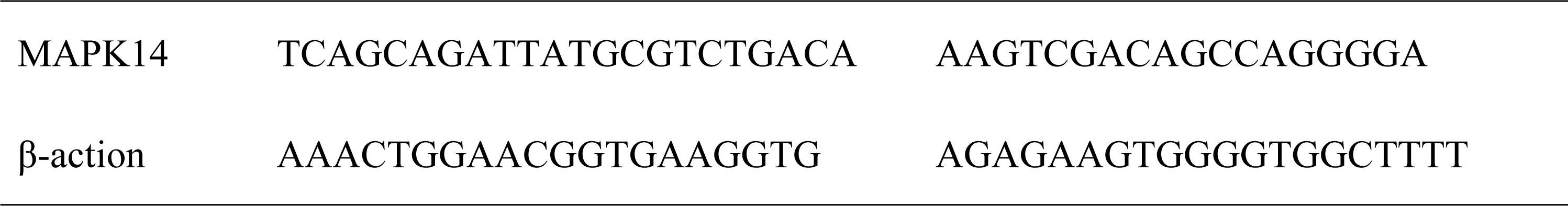
Primer sequences.

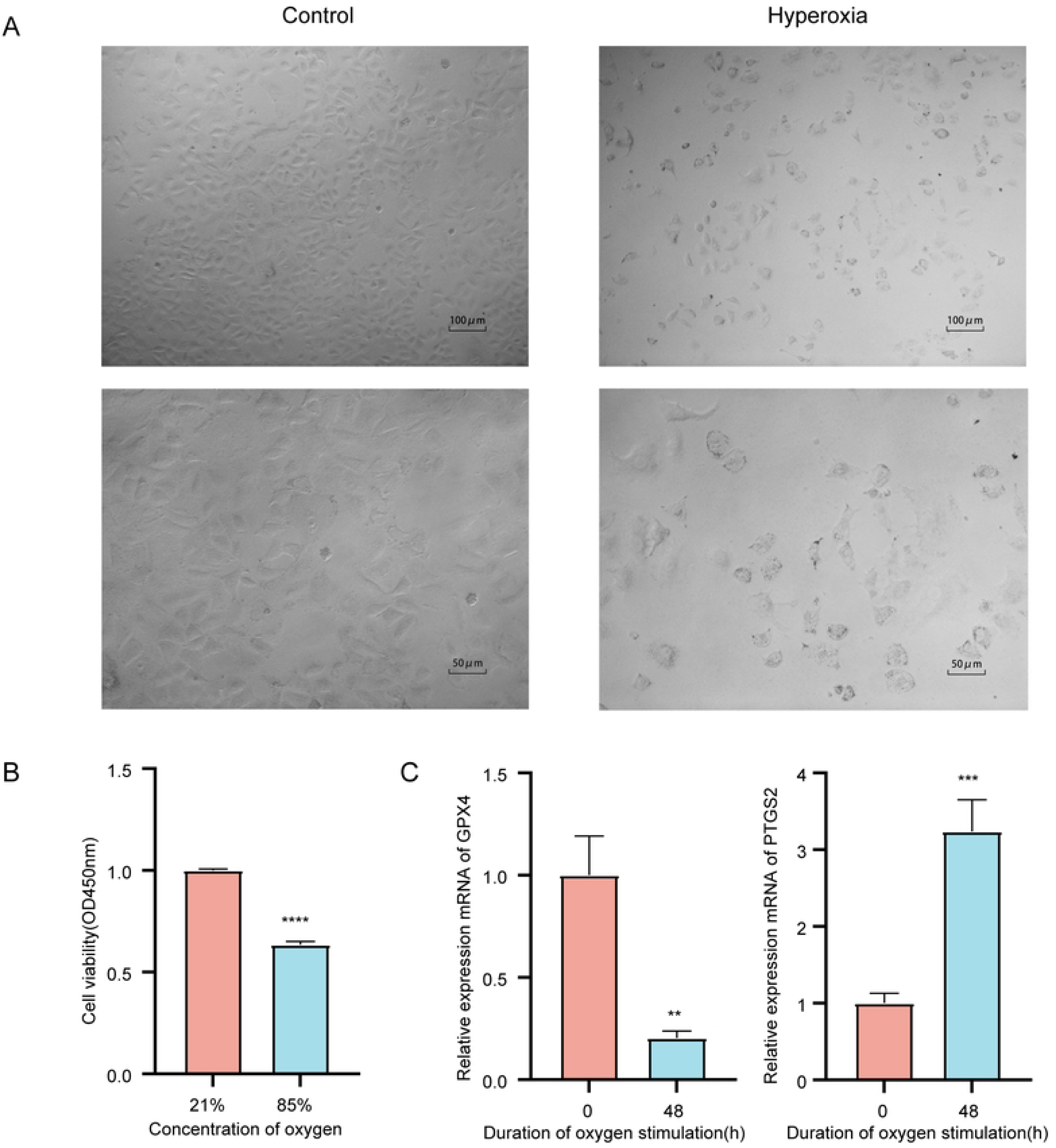

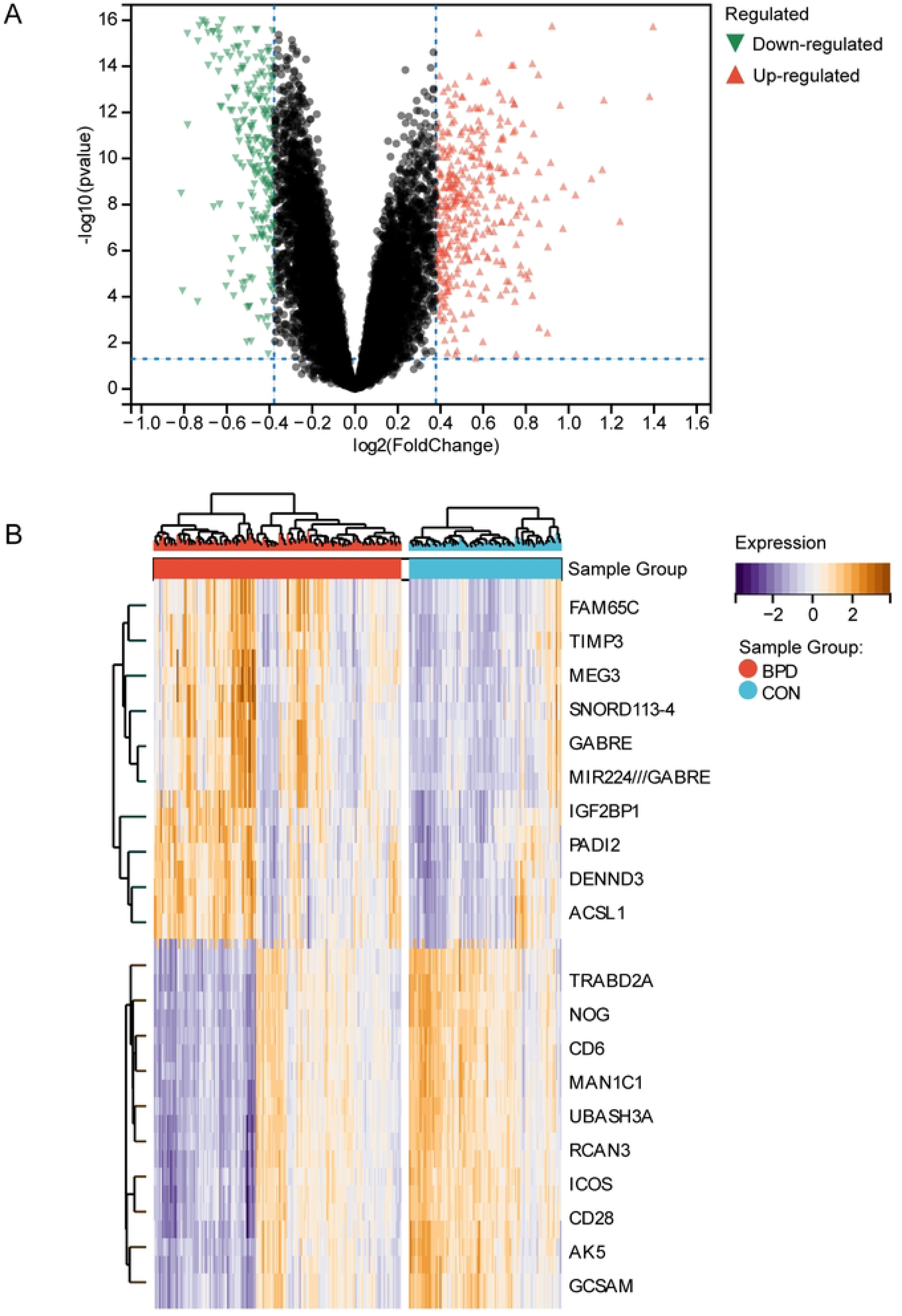

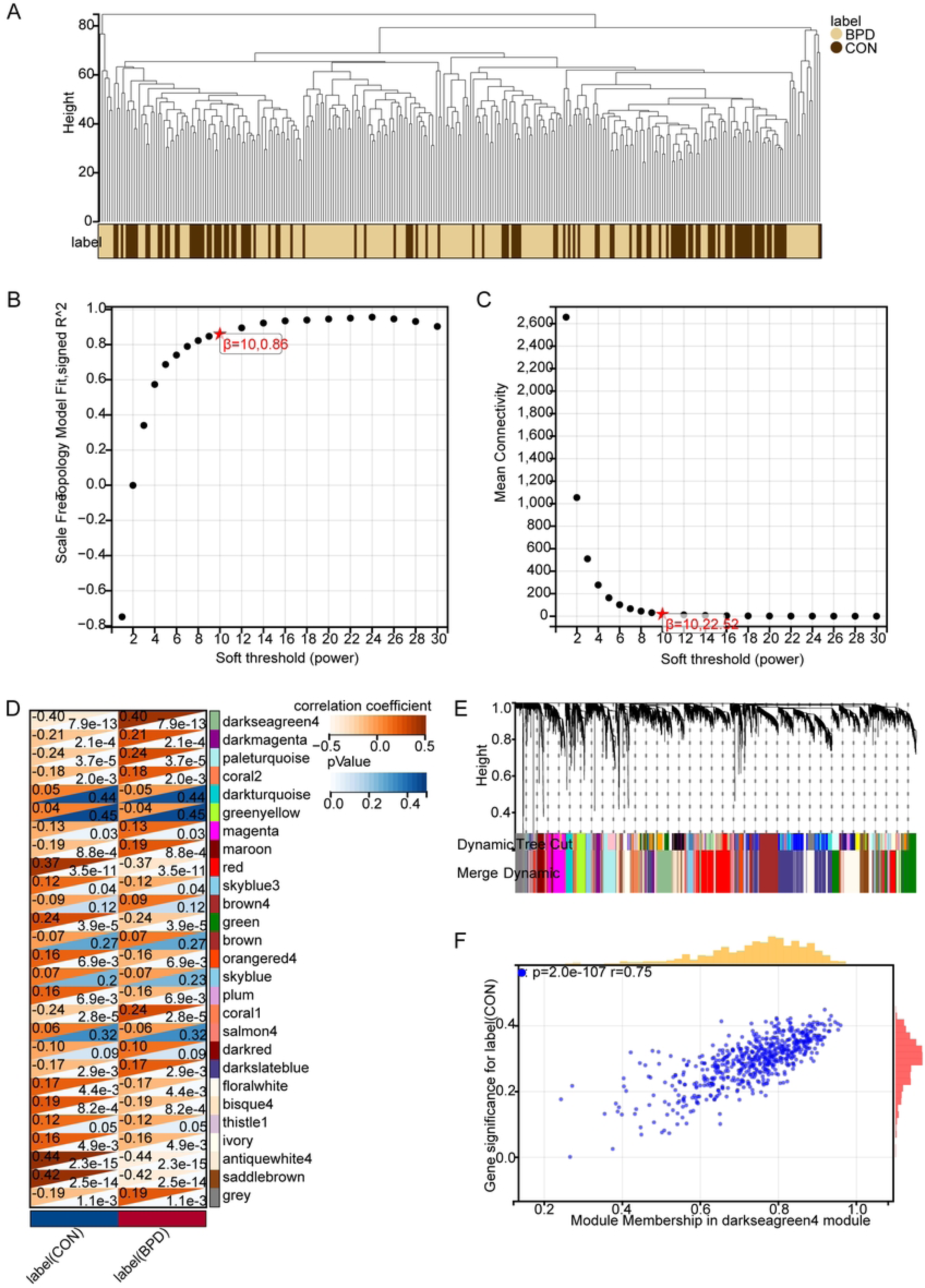

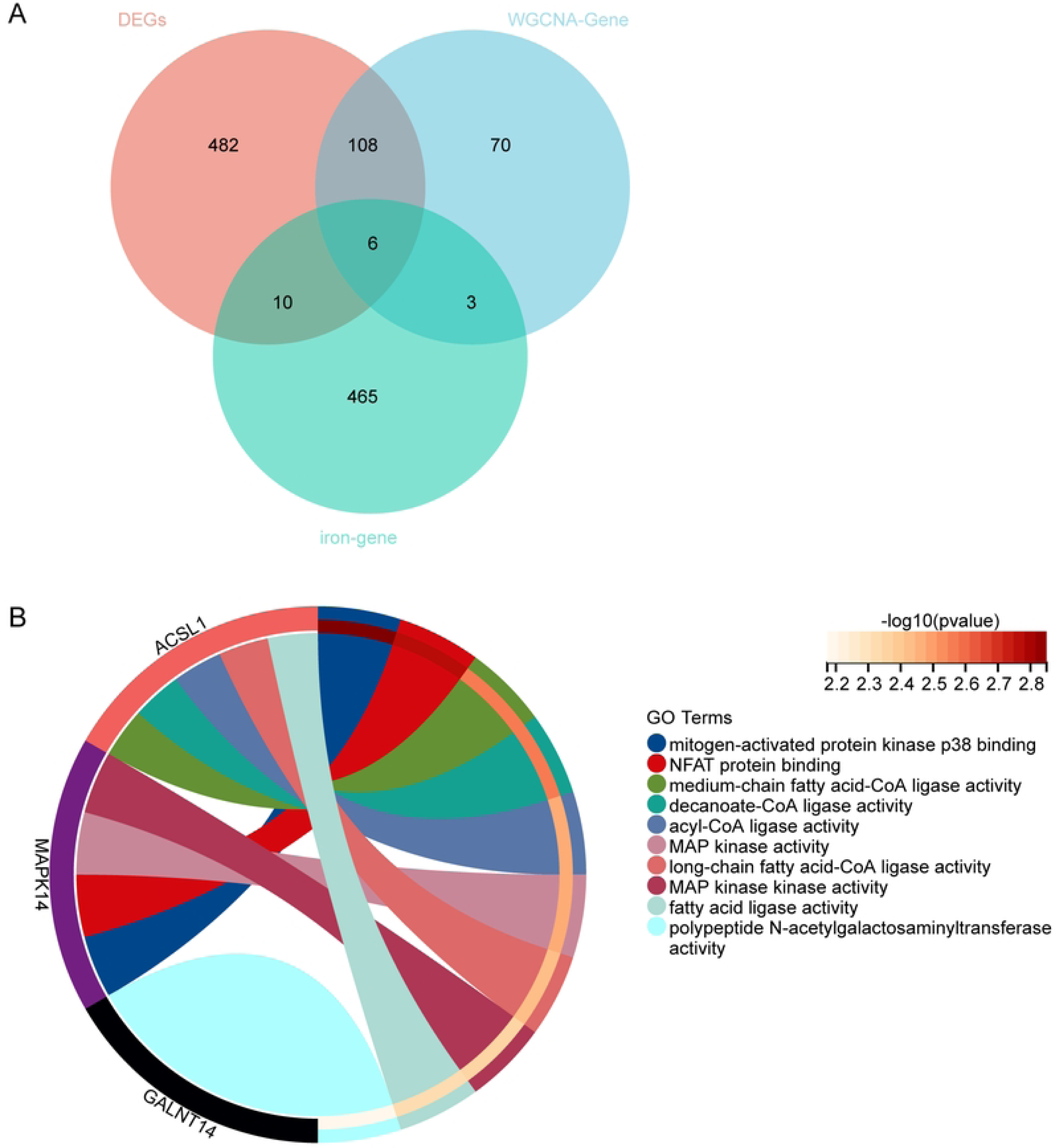

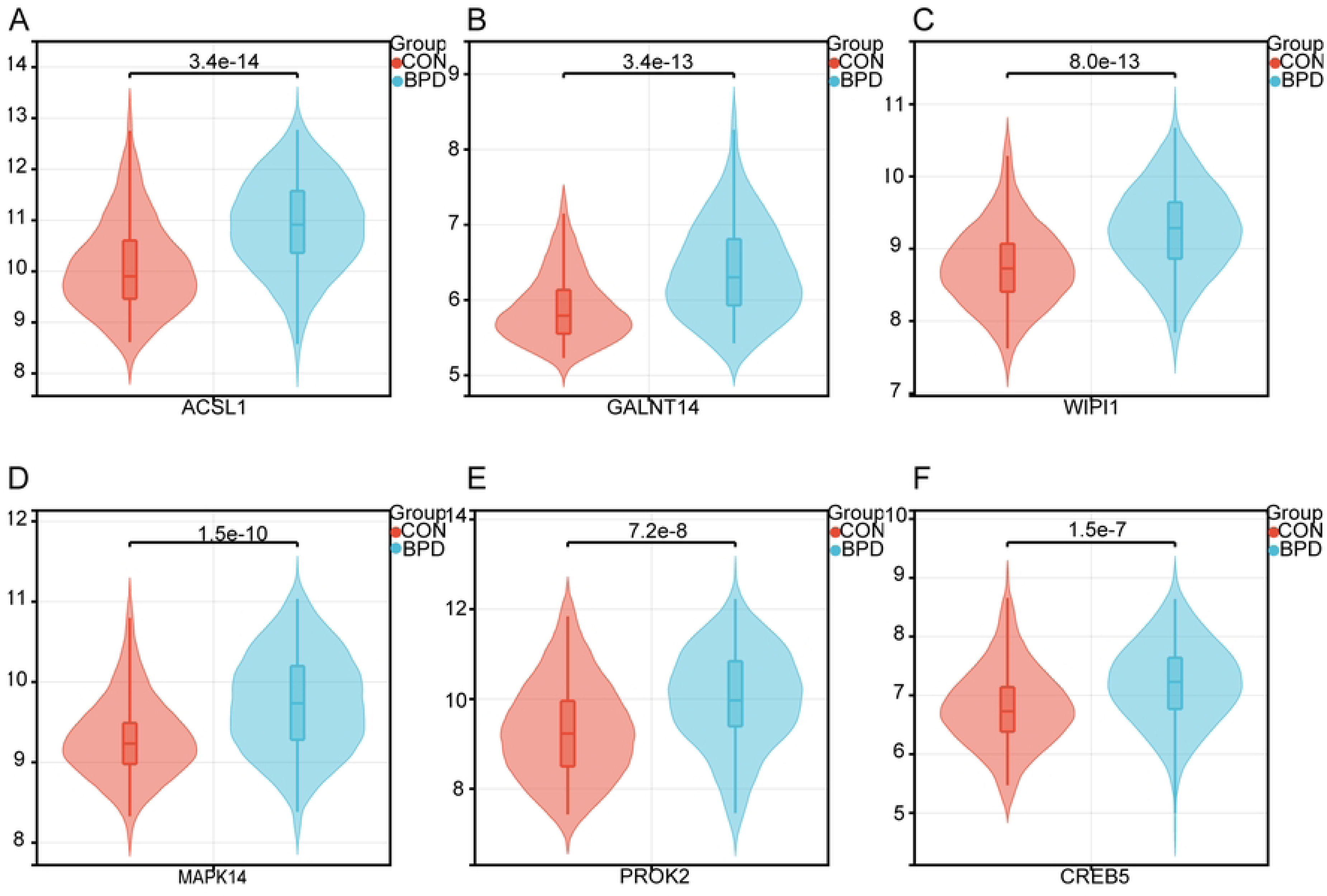

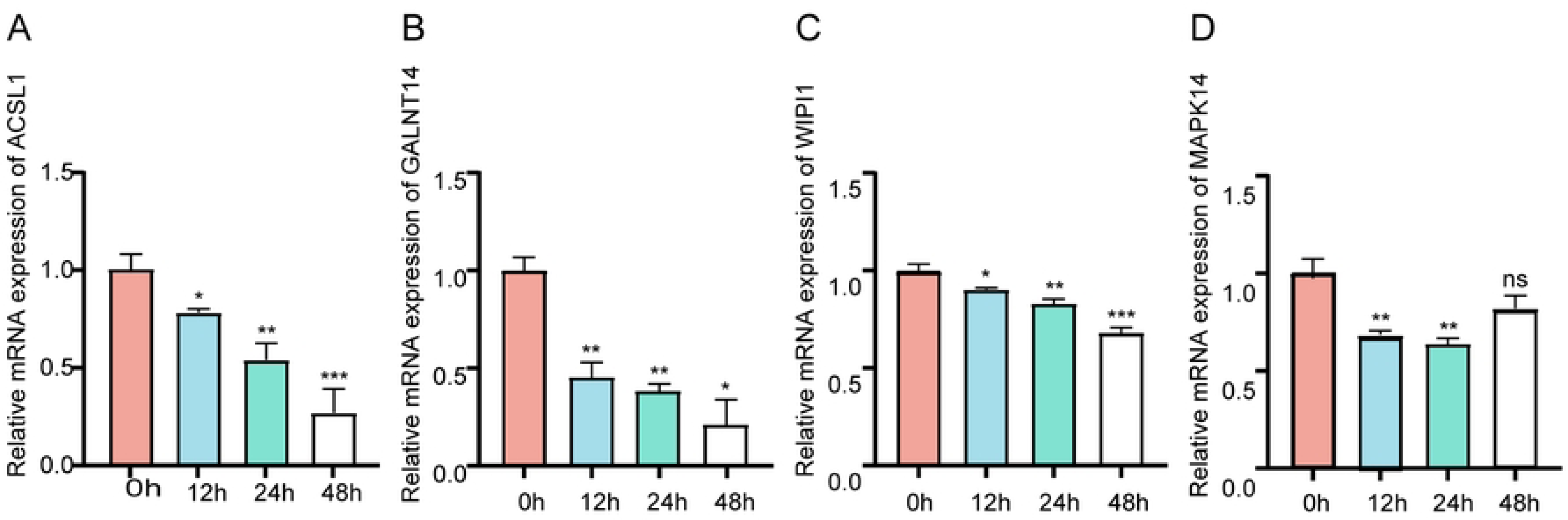

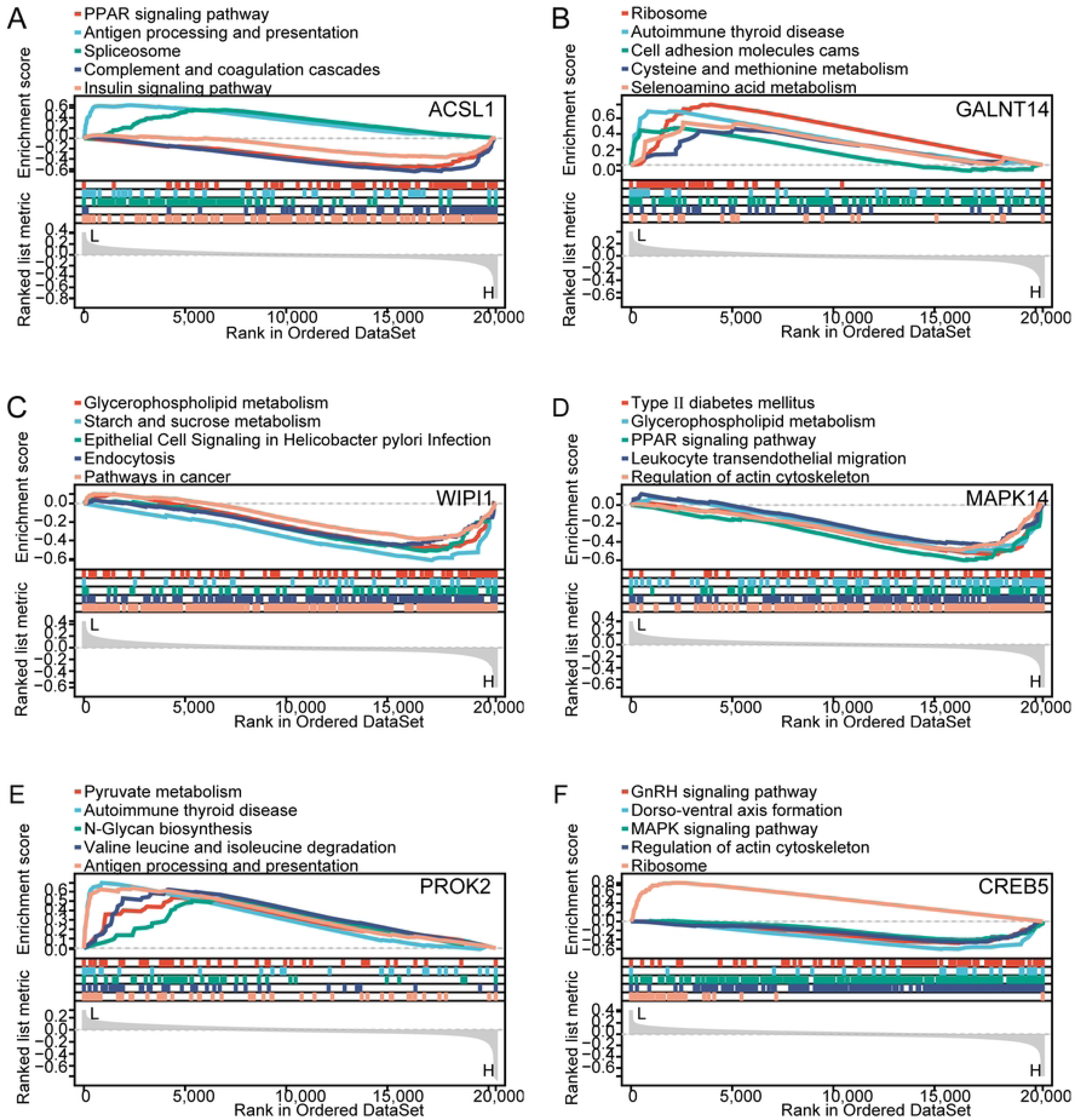

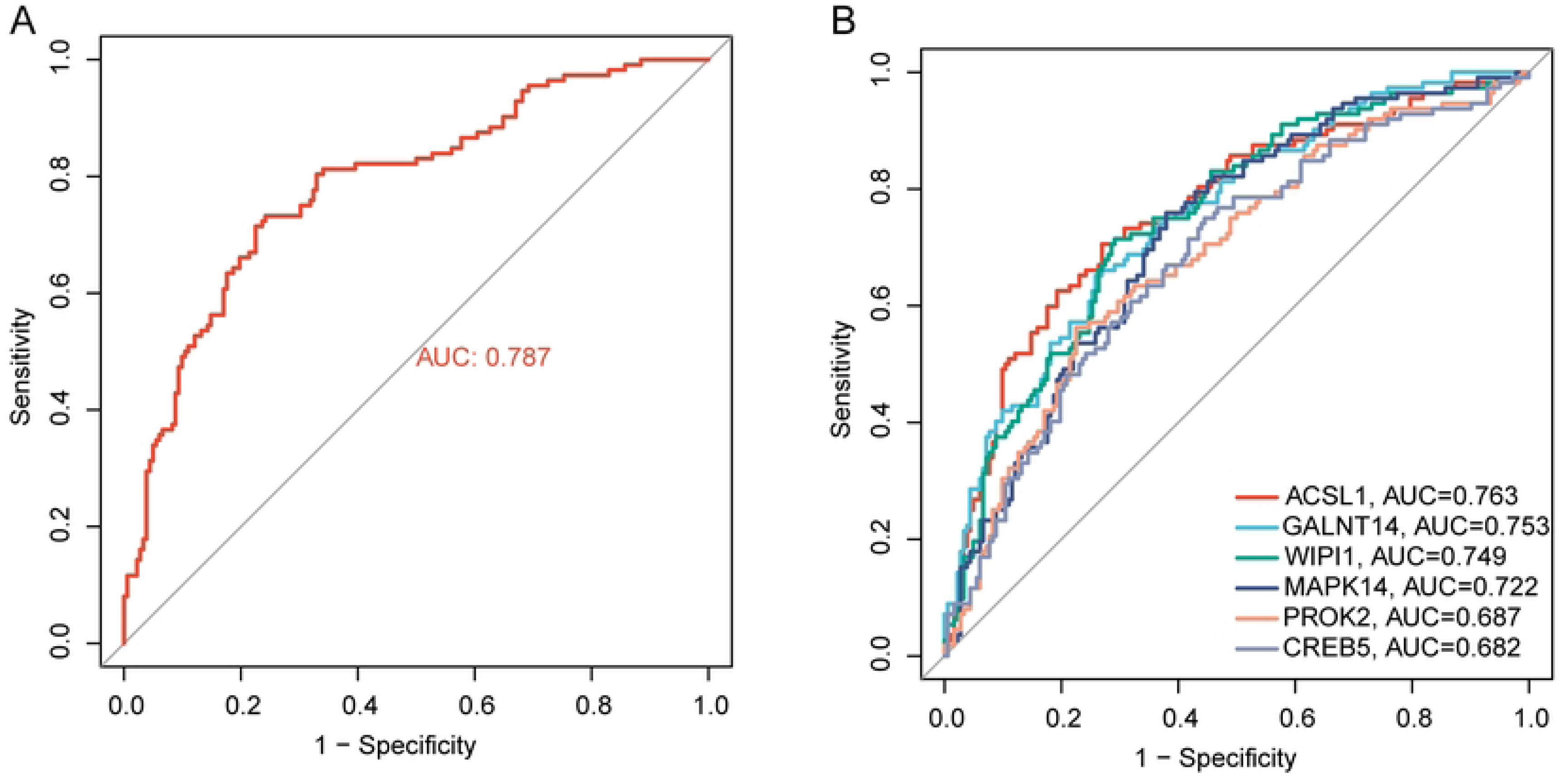

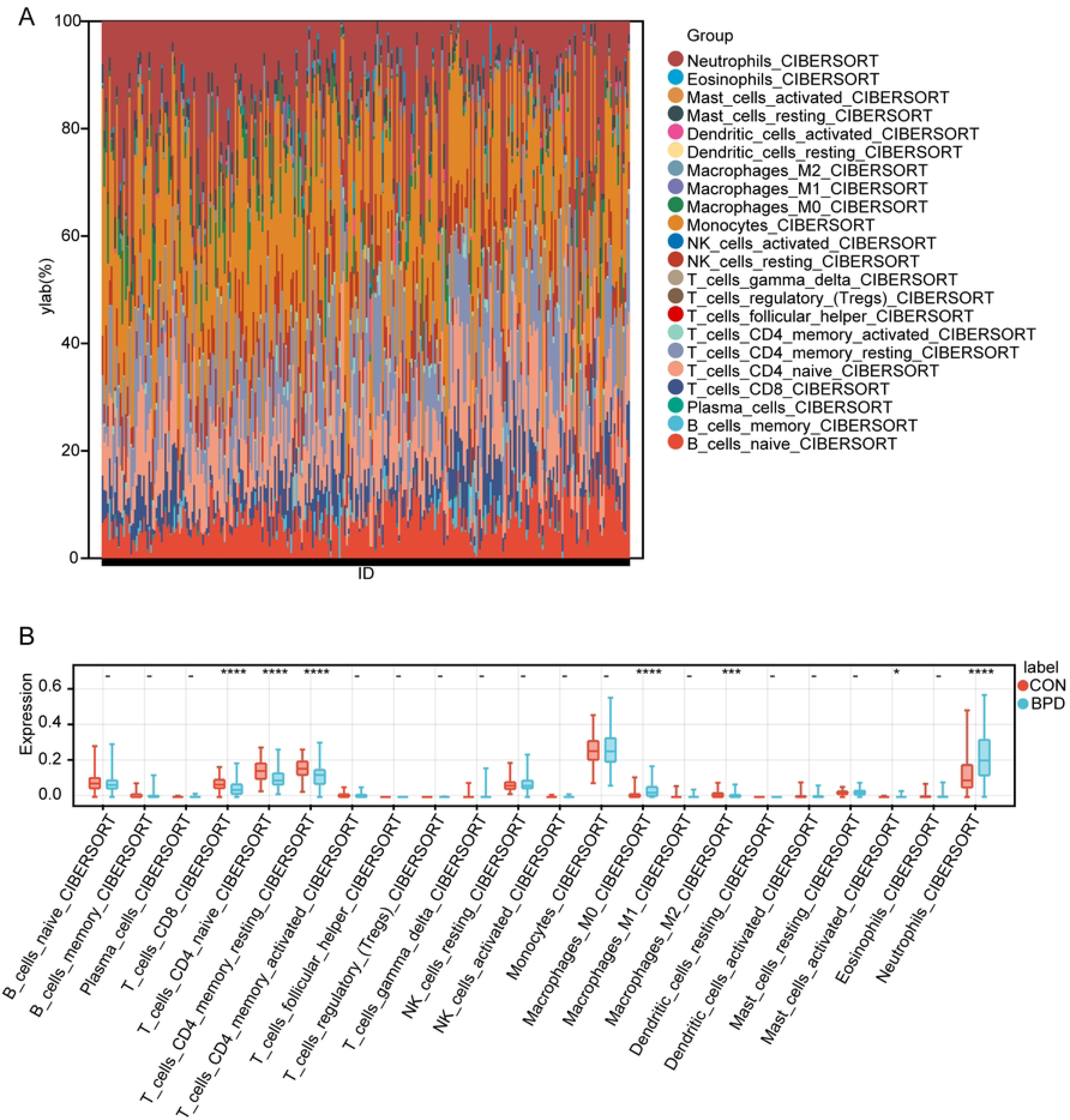

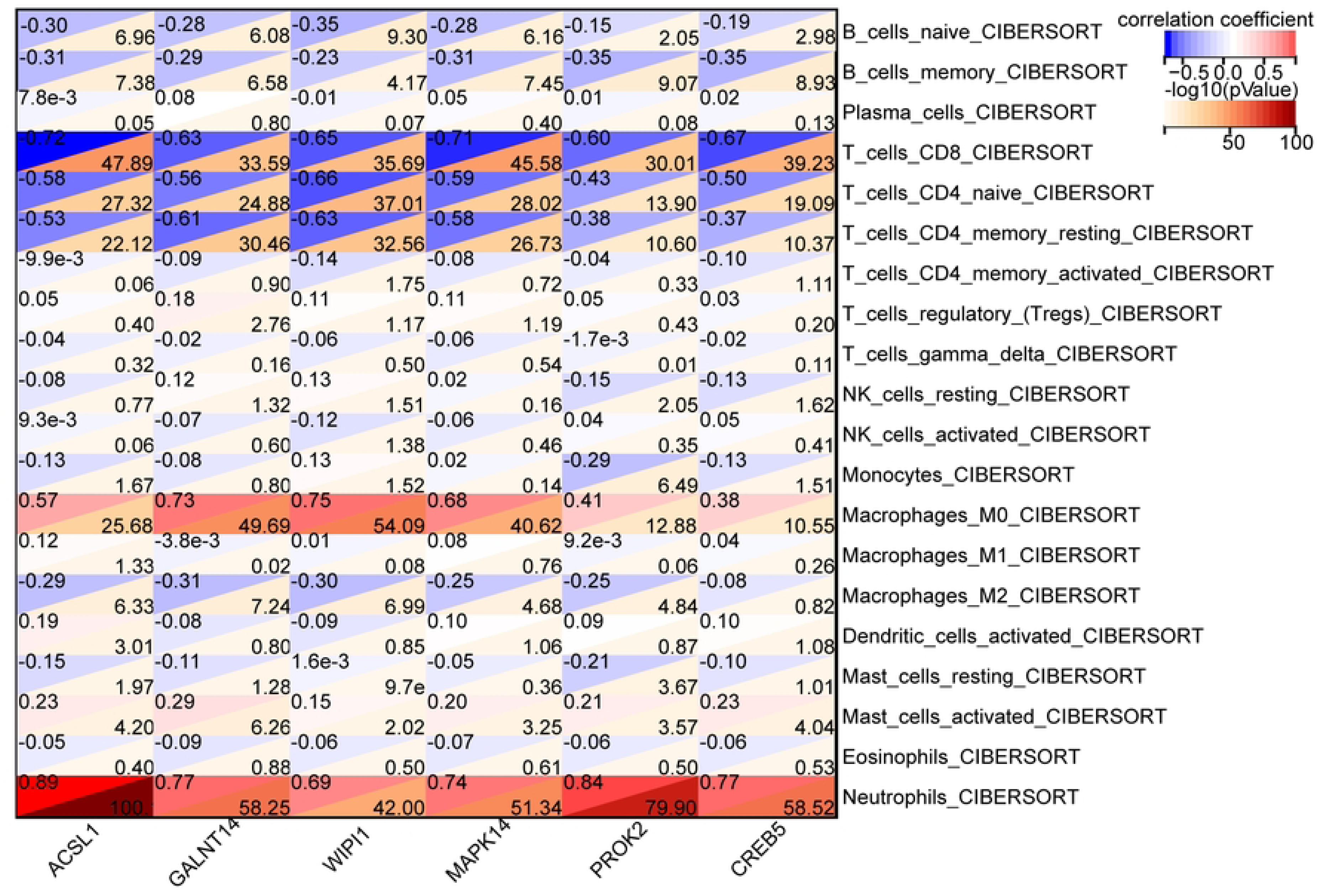

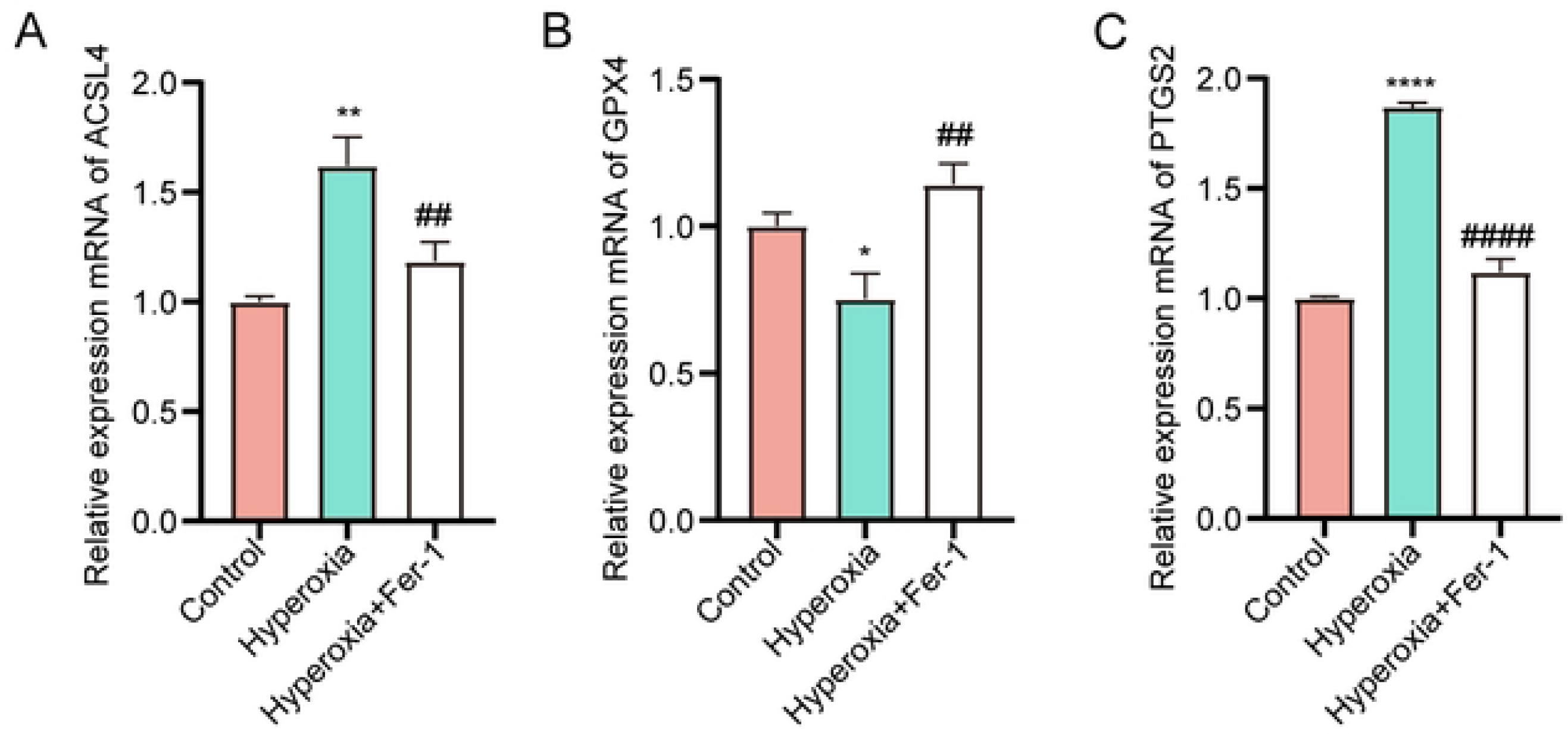

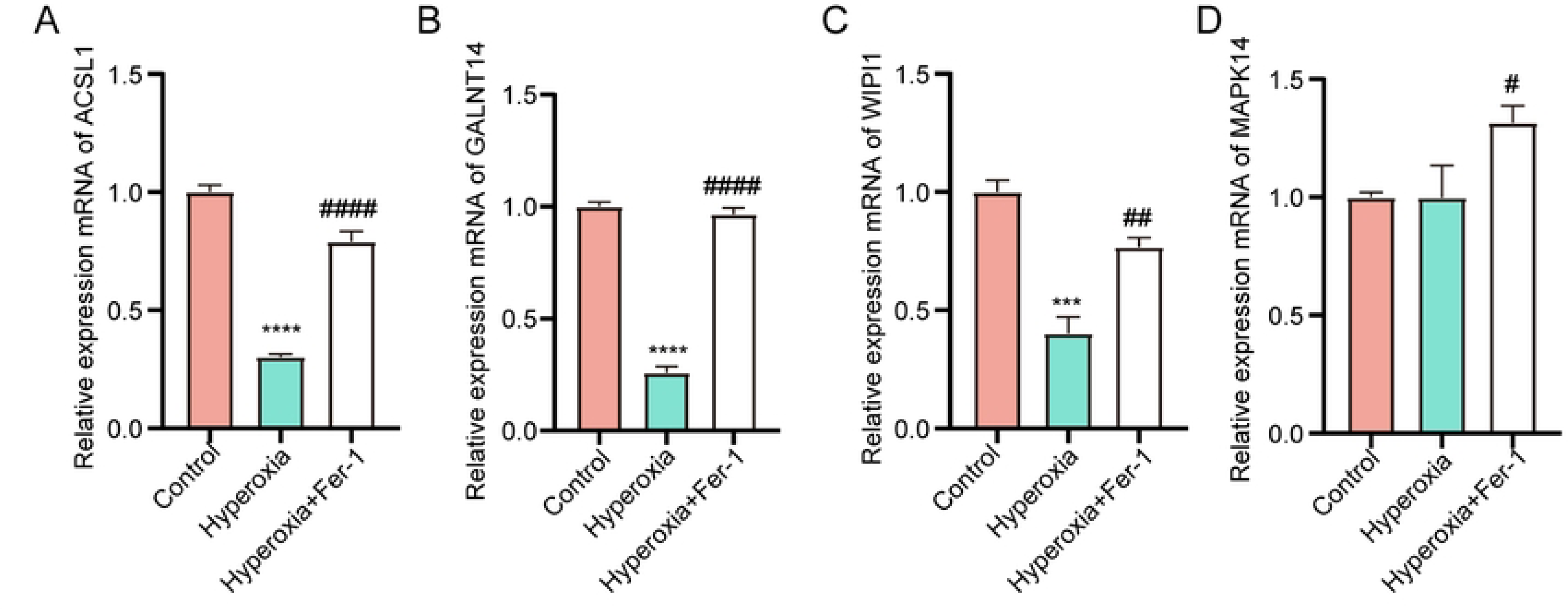

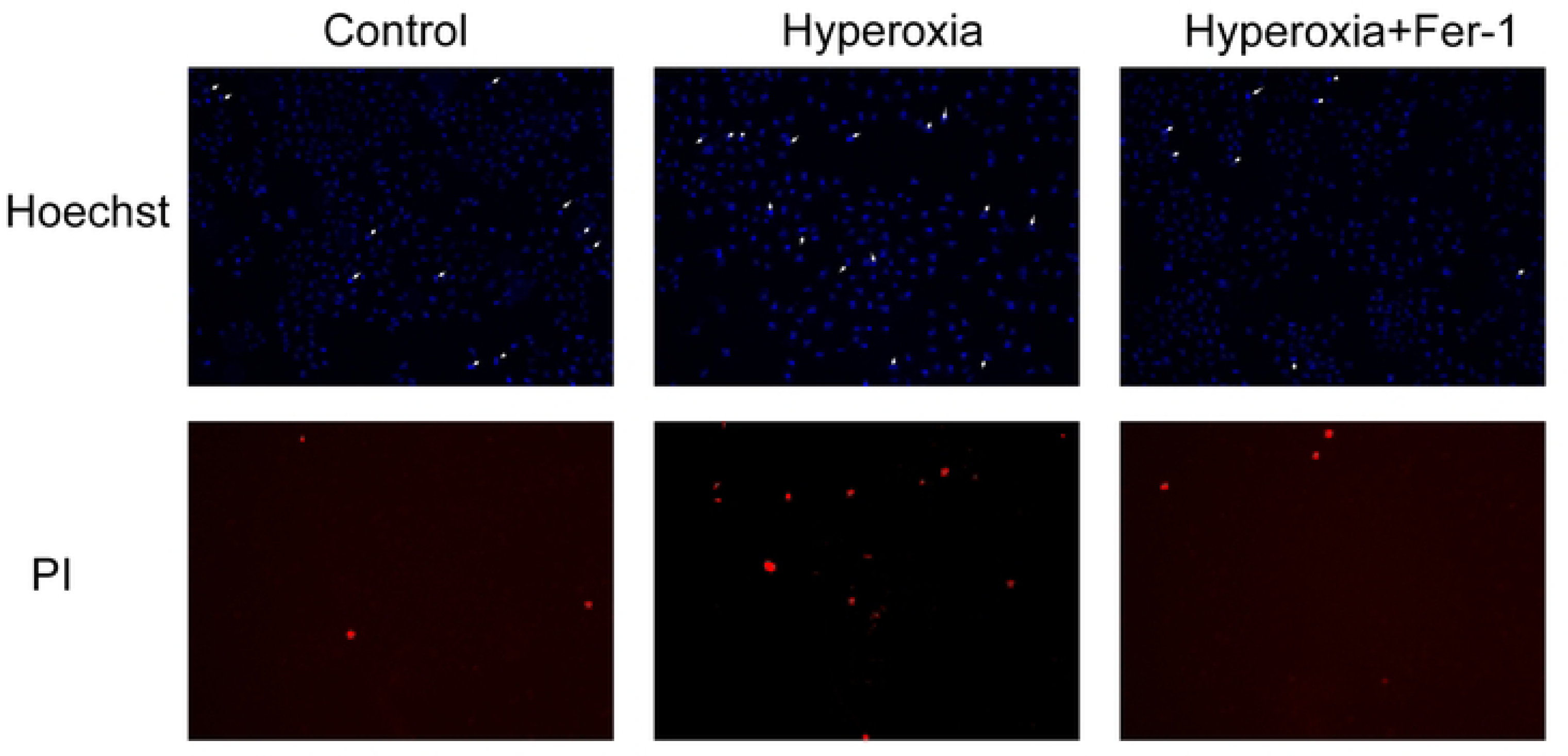

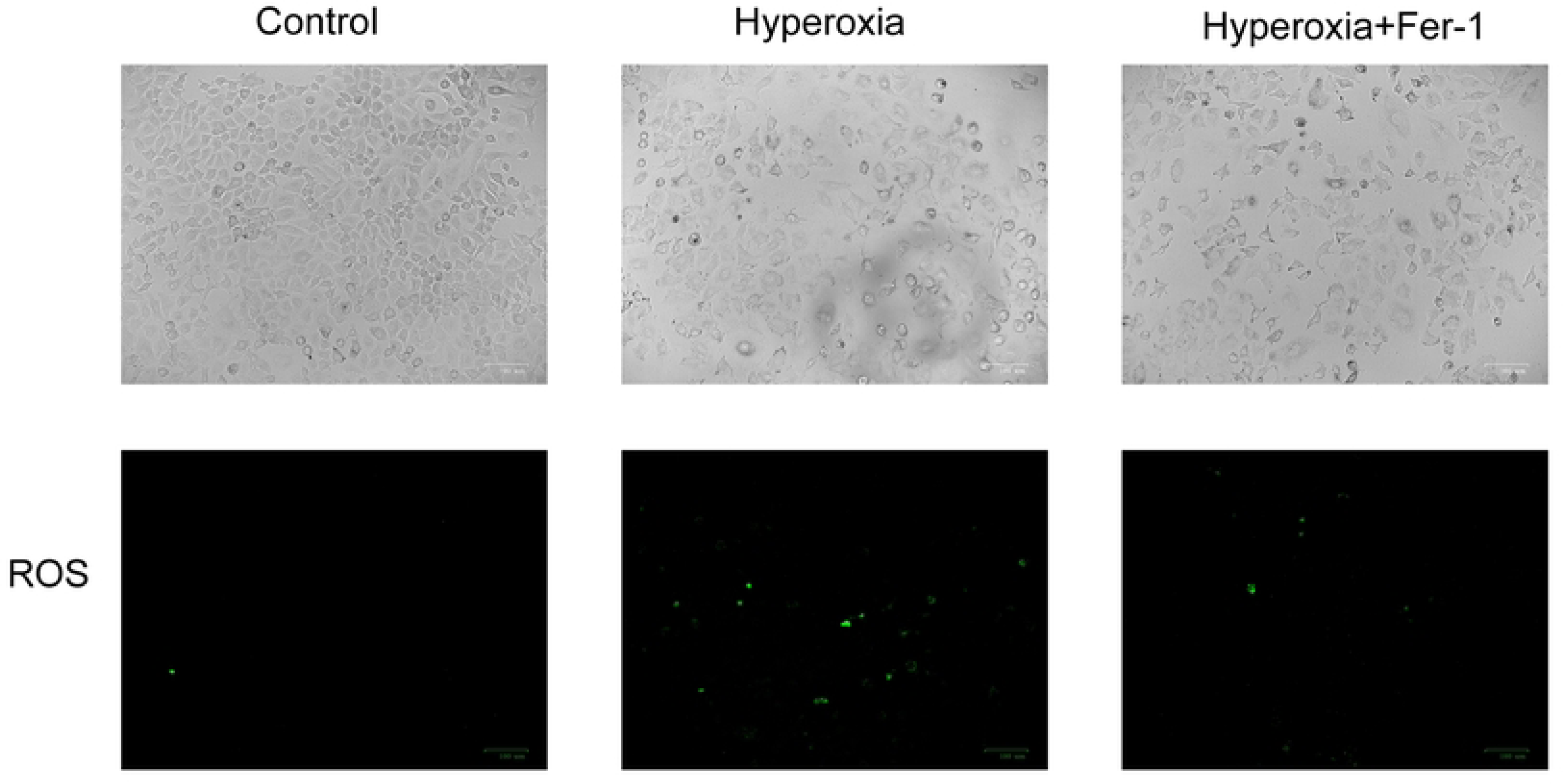

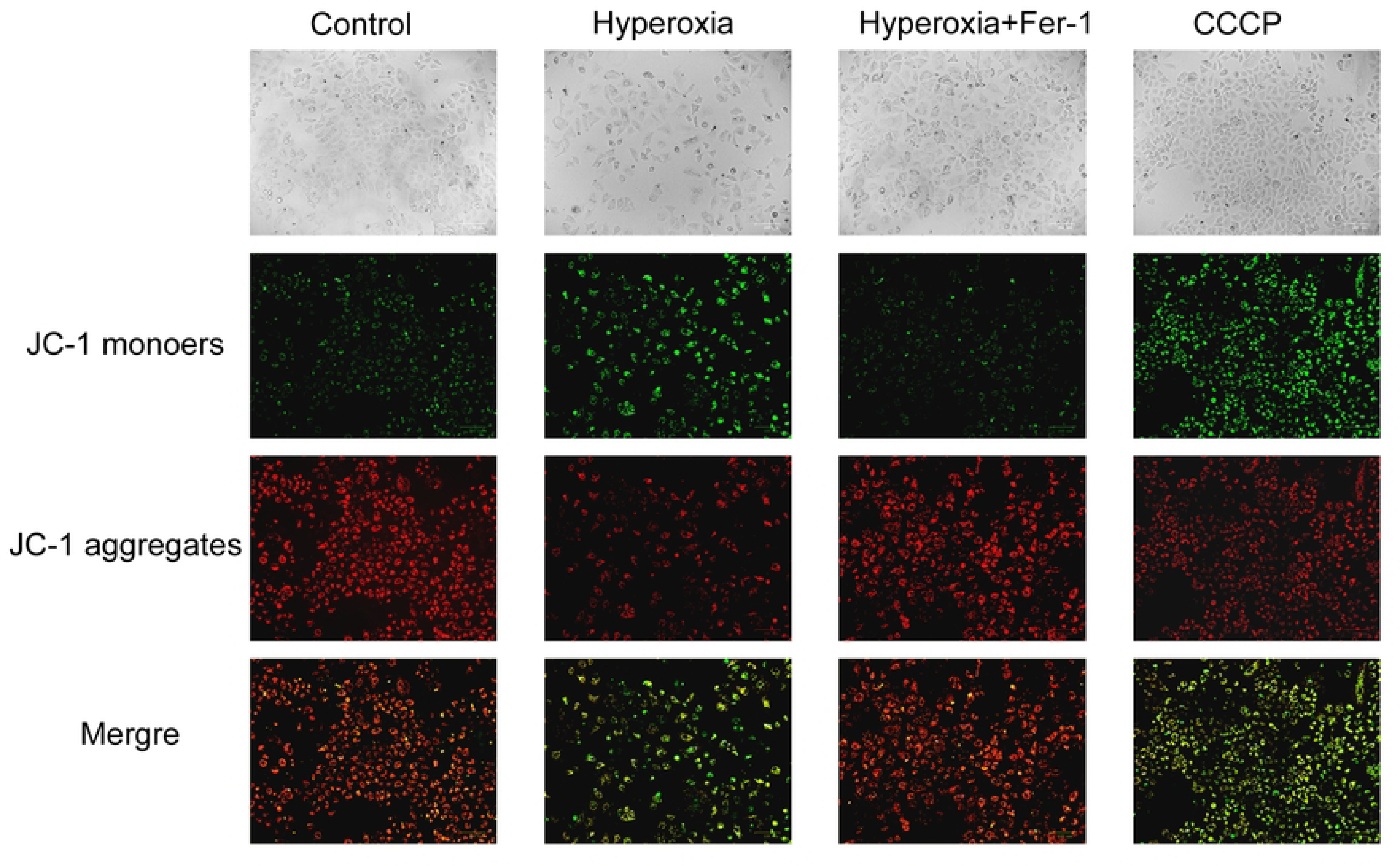

